# Adherence to and enforcement of non-pharmaceutical interventions (NPIs) for COVID-19 prevention in Nigeria, Rwanda, and Zambia: A mixed-methods analysis

**DOI:** 10.1101/2022.03.29.22273120

**Authors:** Hiwote Solomon, Donald M. Thea, Sandro Galea, Lora L. Sabin, Daniel R. Lucey, Davidson H. Hamer

## Abstract

**Introduction:** In the early parts of the COVID-19 pandemic, non-pharmaceutical interventions (NPIs) were implemented worldwide, including in sub-Saharan Africa, to prevent and control SARS-CoV-2 transmission. This mixed-methods study examines adherence to and enforcement of NPIs implemented to curb COVID-19 in Nigeria, Rwanda, and Zambia, leading up to the 10,000^th^ case of laboratory-confirmed COVID-19 in each country. Additionally, we aim to evaluate the relationship between levels and changes of NPIs over time and changes in COVID-19 cases and deaths.

**Methods:** This mixed-methods analysis utilized semi-structured interviews and a quantitative dataset constructed using multiple open data sources, including the Oxford COVID-19 Government Response Tracker. To understand potential barriers and facilitators in implementing and enforcing NPIs qualitative data were collected from those involved in the COVID-19 response and analyzed using NVivo. Quantitative results were analyzed using descriptive statistics, plots, ANOVA, and post hoc Tukey.

**Results:** Individual indicator scores varied with the COVID-19 response in all three countries. Nigeria had sustained levels of strict measures for containment and closure NPIs, while in Rwanda there was substantial variation in NPI score as it transitioned through the different case windows for the same measures. Zambia implemented moderate stringency throughout the pandemic using gathering restrictions and business/school closure measures but maintained low levels of strictness for other containment and closure measures. Rwanda had far more consistent and stringent measures compared to Nigeria and Zambia. Rwanda’s success in implementing COVID-related measures was partly due to strong enforcement and having a population that generally obeys its government.

**Conclusion:** Various forces either facilitated or hindered adherence and compliance to COVID-19 control measures. This research highlights important lessons, including the need to engage communities early and create buy-in, as well as the need for preparation to ensure that response efforts are proactive rather than reactive when faced with an emergency.

## Introduction

In a pandemic, non-pharmaceutical interventions (NPIs) are crucial in curbing disease spread, especially in the absence of vaccines and other pharmaceutical interventions (1). It is widely accepted in public health that early interventions are an important step to halting the progression of a new communicable disease threat (2). Prior to the COVID-19 pandemic, the impact of NPIs has been largely studied in controlling influenza outbreaks, including the 1918/1919 influenza pandemic (1,3). NPIs include actions that can be taken by individuals and the larger community. These include frequent hand washing, covering coughs and sneezes, isolating sick persons, contact tracing, quarantining exposed persons, and physical/social distancing measures for the general population (1,4,5). The latter includes containment strategies such as the closing of schools and workplaces, restricting public gatherings, curfews, quarantine, and maximizing telework (when applicable) (5–8).

The success of NPIs is dependent on enforcement, political governments, and citizens’ willingness to comply with the measures. Until recently, the effectiveness of NPIs has not been tested systematically. Early measures of efficacy were explored mainly through the use of mathematical models, while few published studies have reviewed the historical evidence on NPI adoption during past pandemics (1,3,9). A small number of studies have explored the impact of public health interventions and NPIs during the 1918-1919 influenza pandemic, in which NPIs played a critical role in delaying the temporal effects of the pandemic, in addition to reducing the overall and peak attack rate and the number of cumulative deaths (3,9). How successful NPIs were in limiting disease spread in Africa, especially in the first year of the COVID-19 pandemic has been underexplored in the literature. The degree of implementation and the impact of these NPIs during the COVID-19 pandemic has largely been studied in high-income countries, and there has been limited data in the literature focused on low- and middle-income countries, especially in Africa (10–15).

In the absence of treatment beyond supportive care and vaccination for the early parts of the COVID-19 pandemic, NPIs were implemented across the world to prevent and control the transmission and spread of SARS-CoV-2. By early March 2020, several countries in Africa (affected and unaffected by COVID-19) began mobilizing in response to the pandemic. This included prompt case identification, information campaigns to sensitize citizens, and building laboratory capacity (16,17). Some countries relied on innovative strategies such as using locally produced cloth masks, soaps, and hand sanitizers, developing inexpensive diagnostic tests, testing pooled COVID-19 samples, and using drones to transport test kits and samples to and from hard-to-reach areas (18–20). By the end of March 2020, many African Union Member States had imposed travel bans on flights arriving from certain Asian and European countries (20). In the following two months, almost two-thirds of African Union Member States had closed their borders to all international travelers, except for cargo, freight, and expatriation of foreign nationals (19,20). Fifteen countries, including Nigeria and Rwanda, implemented border closures before any COVID-19 cases were confirmed (21). Other NPIs, such as restrictions of movement and public gathering, and closure of schools and workplaces were also implemented across the region.

The present study focuses on three countries in Africa: Nigeria in West Africa, Rwanda in East Africa, and Zambia in southern Africa. These countries were selected to provide variety in geopolitical structures, population size, region, and World Bank income classification. This mixed-methods study aims to examine adherence and enforcement of NPIs implemented to curb COVID-19 in Nigeria, Rwanda, and Zambia, leading up to the 10,000^th^ recorded case of COVID-19 in each country. Additionally, we aim to broadly evaluate the relationship among the levels/changes in NPIs implemented and changes in COVID-19 cases and deaths.

## Methods and Materials

### Study design

This mixed-methods study utilized a mix of semi-structured interviews and a quantitative dataset constructed using multiple open data sources.

### Qualitative method

To understand the potential barriers and facilitators in implementing and enforcing NPIs and how other epidemics within the countries may have affected compliance in NPIs, qualitative data were collected from decision-makers and experts involved in the COVID-19 response using key informant interviews (KIIs). KIIs were conducted with officials of Ministries of Health, Africa CDC Regional Collaborating Centers, and WHO African Regional Office (AFRO). Recruitment utilized purposive and convenience sampling, including a snowball sampling approach. All KIIs provided verbal consent before the start of the interview. Each KII took on average 30 minutes to complete using Zoom. The audio recordings were downloaded from Zoom and then immediately uploaded to a secure database and deleted from the computer. Transcription took place upon completion of the interviews. Data were collected and analyzed using a grounded theory approach (22). Thematic analysis using inductive coding was used to systematically extract key themes in an iterative process as they emerged through the analysis process. An iterative process was used to develop a comprehensive codebook. During the initial coding phase, first-order codes were developed, while secondary coding allowed for the grouping of first-order codes into themes. Quotes attributed to specific themes were extracted. Both coding and analysis were conducted using NVivo Release 1.6 Mac Edition (23).

### Quantitative Methods

A time-series dataset was constructed using multiple open data sources. Observations for each variable were recorded daily beginning on January 1^st^, 2020, for each of the three countries (Nigeria, Rwanda, and Zambia) and ending on the date when each country surpassed its 10,000^th^ case. While most studies exploring the effect of NPIs have focused on 100 cases as the outbreak threshold, this study uses four case windows to gain a broader picture: no cases (W_0_), 1-100 cases (W_1_), >101-1,000 cases (W_2_), and >1,001-10,000+ cases (W_3_)). New and cumulative cases during this period were obtained from the COVID-19 data repository by the Center for Systems Science and Engineering at Johns Hopkins University (24). In addition to COVID-19 case and death aggregates, the dataset also includes variables from the University of Oxford COVID-19 Government Response Tracker (OxCGRT), which collects publicly available information on 20 indicators in different areas, such as containment policies, economic policies, and health policies in more than 150 countries since January 2020 (25). Data are collected in real-time and from publicly available sources in each country. The OxCGRT dataset utilized in this paper was downloaded in March 2021.

This paper focuses on two policy indices calculated by the OxCGRT, the stringency index (SI) and the containment and health index (CHI). Table 1 shows which variables were included in each index. The SI records the strictness of closure and containment policies using 9 indicators, which include lockdown policies, primarily aimed at restricting certain behaviors, while the CHI includes all the variables from the stringency index and additional variables focused on mitigating the health consequences of COVID-19 (e.g., testing, use of facial coverings outside the home, and contact tracing) (25). The methods and calculation of indices are described elsewhere by Hale et al (25). Broadly, both indices aggregate the data of individual policy measures into a single number, between 0 to 100, that reflects the level of a government’s response along certain dimensions to measure the indicators upon which a government has acted, and to what degree.

**Table 1.** Non-pharmaceutical policy variables used in OxCGRT index calculation.

| Variable | Description | SI | CHI |
| --- | --- | --- | --- |
| C1 | Record closings of schools and universities | X | X |
| C2 | Record closings of workplaces | X | X |
| C3 | Record canceling public events | X | X |
| C4 | Record limits on gatherings | X | X |
| C5 | Record closing of public transport | X | X |
| C6 | Record orders to "shelter-in-place" and otherwise confine to the home | X | X |
| C7 | Record restrictions on internal movement between cities/regions | X | X |
| C8 | Record restrictions on international travel (this records policy for foreign travelers, not citizens) | X | X |
| H1 | Record presence of public info campaigns | X | X |
| H2 | Record government policy on who has access to testing<br>Note: this records policies about testing for current infection (PCR tests) not testing for immunity (antibody test) |  | X |
| H3 | Record government policy on contact tracing after a positive diagnosis<br>Note: Policies include only those that would identify all people potentially exposed to COVID-19 |  | x |
| H6 | Record policies on the use of facial coverings outside the home |  | x |
| H7 | Record policies for protecting elderly people (as defined locally) in Long Term Care Facilities and/or the community and home setting |  | x |

To examine the degree of implementation of NPIs relative to the four case windows, we used plots and descriptive statistics (means and standard deviations) stratified by the case windows. Indices were compared across case windows using analysis of variance (ANOVA) models with post-hoc comparisons (p-values adjusted using Tukey’s method). All analyses were performed using SAS statistical software version 9.4 (SAS Institute Inc., Cary, NC) (26).

### Ethical Considerations

IRB exemption was granted on May 5, 2021, by the Boston University Medical Center Institutional Review Board (IRB number: H-41329). All key informant interviewees were informed that participation was voluntary. All participants provided verbal consent to being recorded before the start of the interview.

## Results

### Quantitative findings

It took Rwanda less time to surpass the first 100 cases (21 days) compared to Nigeria (30 days) and Zambia (43 days); however, it took Rwanda substantially longer to surpass 10,000 cases (379 days) than Nigeria and Zambia (152 and 232 days, respectively). All three countries experienced an exponential rise in cases after surpassing 1,000 cases of COVID-19.

An examination of SI scores from January 1^st^, 2021, until each country surpassed 10,000 cases, reveals several important differences across countries (Tables 2a-2c). In Rwanda, the average score of the SI from January 1^st^, 2020, until it surpassed 10,000 cases was 61.1 out of 100 (SD = 26.6, median=73.2) indicating sustained moderate stringency on measures. In contrast, Nigeria and Zambia had average scores of 42.1 (SD = 37.1, median=11.1) and 34.3 (SD = 22.3, median=43.5), respectively. In Nigeria, a Tukey post hoc test shows the mean SI scores in case windows W_2_ (SI=83.0) and W_3_ (SI=84.6) were statistically significantly different (p<0.0001) when compared to W_0_ (SI=6.2). However, there was no difference between the mean SI scores between W_2_ and W_3_ (p=0.85), indicating stringency scores stayed relatively around the same level after the 100^th^ case. In contrast, in Zambia, the mean SI scores in case windows W_1_ (SI= 49.5), W_2_ (SI=51.2), and W_3_ (SI=47.4) were significantly different (p<0.0001) than W_0_ (SI=5.6), indicating that Zambia’s stringency levels varied post-identification of the index case compared to pre-identification. However, there was no difference between the mean SI scores across case windows W_1_, W_2_, and W_3_, suggesting that the stringency levels in how the measures were implemented stayed relatively stable despite cases increasing. In Rwanda, mean SI scores in W_1_ (SI=73.2) and W_3_ (SI=70.3) were statistically significantly different (p<0.05) when compared to W_2_ (SI=79.5). However, there was no difference between the mean SI scores between W_1_ and W_3_ (p=0.53) suggesting that the stringency levels were similar in terms of which measures were implemented during W_1_ and W_3_.

**Table 2a.** One-Way Analysis of Variance of the Stringency Index Score by Case Window, Nigeria.

| Case Window | *n* | Mean | *SD* | Tukey's Post-hoc Comparisons |  |  |  |
| --- | --- | --- | --- | --- | --- | --- | --- |
| | | | | $W_0$ | $W_1$ | $W_2$ | $W_3$ |
| **Pooled** | 152 | 42.1 | 37.1 | - | - | - | - |
| **$W_0$** | 58 | 6.2 | 5.1 |  | <0.0001 | <0.0001 | <0.0001 |
| **$W_1$** | 30 | 22.3 | 16.3 | <0.0001 |  | <0.0001 | <0.0001 |
| **$W_2$** | 26 | 83.0 | 0.6 | <0.0001 | <0.0001 |  | 0.85 |
| **$W_3$** | 38 | 84.6 | 0.6 | <0.0001 | <0.0001 | 0.85 |  |

Note: $W_0$ : 0 cases, $W_1$ : 1-100+ cases, $W_2$ : >101-1,000+ cases, $W_3$ : >1,001-10,000+ cases *n*: number of days, *SD*: standard deviation

**Table 2b.** One-Way Analysis of Variance of the Stringency Index by Case Window, Rwanda.

| Case Window | *n* | Mean | *SD* | Tukey's HSD Comparisons |  |  |  |
| --- | --- | --- | --- | --- | --- | --- | --- |
| | | | | $W_0$ | $W_1$ | $W_2$ | $W_3$ |
| **Pooled** | 379 | 61.1 | 26.6 | - | - | - | - |
| **$W_0$** | 73 | 10.9 | 7.6 |  | <0.0001 | <0.0001 | <0.0001 |
| **$W_1$** | 21 | 73.2 | 25.6 | <0.0001 |  | 0.04 | 0.54 |
| **$W_2$** | 86 | 79.5 | 8.6 | <0.0001 | 0.04 |  | <0.0001 |
| **$W_3$** | 199 | 70.3 | 7.5 | <0.0001 | 0.54 | <0.0001 |  |

Note: $W_0$ : 0 cases, $W_1$ : 1-100+ cases, $W_2$ : >101-1,000+ cases, $W_3$ : >1,001-10,000+ cases *n*: number of days, *SD*: standard deviation

**Table 2c.**
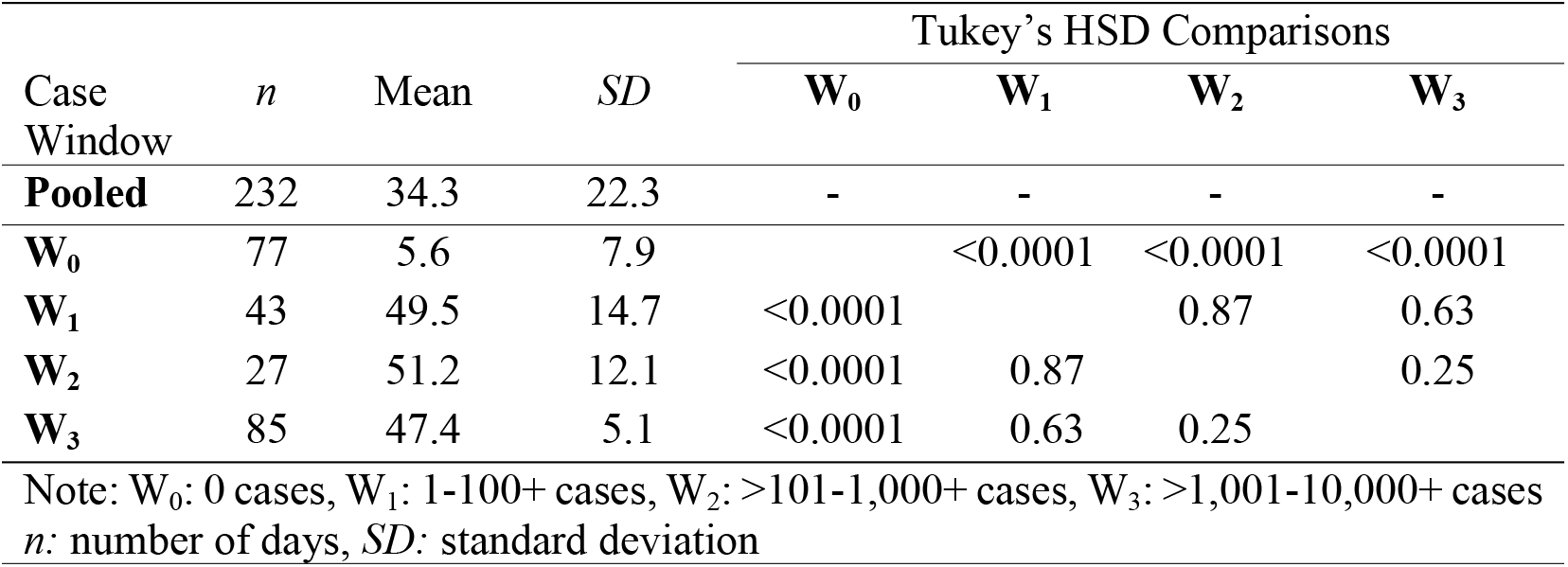
One-Way Analysis of Variance of the Stringency Index by Case Window, Zambia.

There were also major differences between countries when examining the degree of implementation of the CHI which measured all eight containment and closure NPIs and eight health system NPIs. In Rwanda, the overall average score for the CHI was 58.5 (SD = 25.3, median=71.4) (Table 3a). In contrast, the average scores in Nigeria and Zambia were 38.3 (SD = 32.2, median=16.7) and 31.1 (SD = 20.8, median=40.5), indicating lower stringency and levels of implementation compared to Rwanda. When looking at differences in implementation between case windows, there were significant differences between the CHI scores across case windows in Nigeria indicating varying levels of implementation between the case windows. By comparison, in Rwanda, the mean containment and health index scores in case windows W_1_ (CHI=65.3) and W_3_ (CHI=68.7) were significantly different (p<0.001) compared to W_2_ (CHI=74.7). However, there was no difference between the mean CHI scores in W_1_ and W_3_, indicating that the implementation of measures calculated in the CHI were similar during W_1_ and W_3_. Lastly, in Zambia, there was a statistically significant difference between the mean containment and health index scores in case windows W_1_ (CHI=41.7), W_2_ (CHI=47.8), and W_3_ (CHI=45.3). However, there was no difference in the mean containment and health index scores between W_1_ and W_2_ compared to W_3_ (p>0.05), which again indicates similar implementation of the measures calculated in the containment and health index during W_1_, W_2_, and W_3_.

**Table 3a.** One-Way Analysis of Variance of the Containment and Health Index by Case Window, Nigeria.

| Case Window | *n* | Mean | *SD* | Tukey's Post-hoc Comparisons |  |  |  |
| --- | --- | --- | --- | --- | --- | --- | --- |
| | | | | $W_0$ | $W_1$ | $W_2$ | $W_3$ |
| **Pooled** | 152 | 38.3 | 32.2 | - | - | - | - |
| **$W_0$** | 58 | 5.1 | 4.4 |  | <0.0001 | <0.0001 | <0.0001 |
| **$W_1$** | 30 | 26.5 | 13.8 | <0.0001 |  | <0.0001 | <0.0001 |
| **$W_2$** | 26 | 70.5 | 1.0 | <0.0001 | <0.0001 |  | 0.0065 |
| **$W_3$** | 38 | 76.1 | 0.4 | <0.0001 | <0.0001 | 0.0065 |  |

Note: $W_0$ : 0 cases, $W_1$ : 1-100+ cases, $W_2$ : >101-1,000+ cases, $W_3$ : >1,001-10,000+ cases *n*: number of days, *SD*: standard deviation

**Table 3b.**
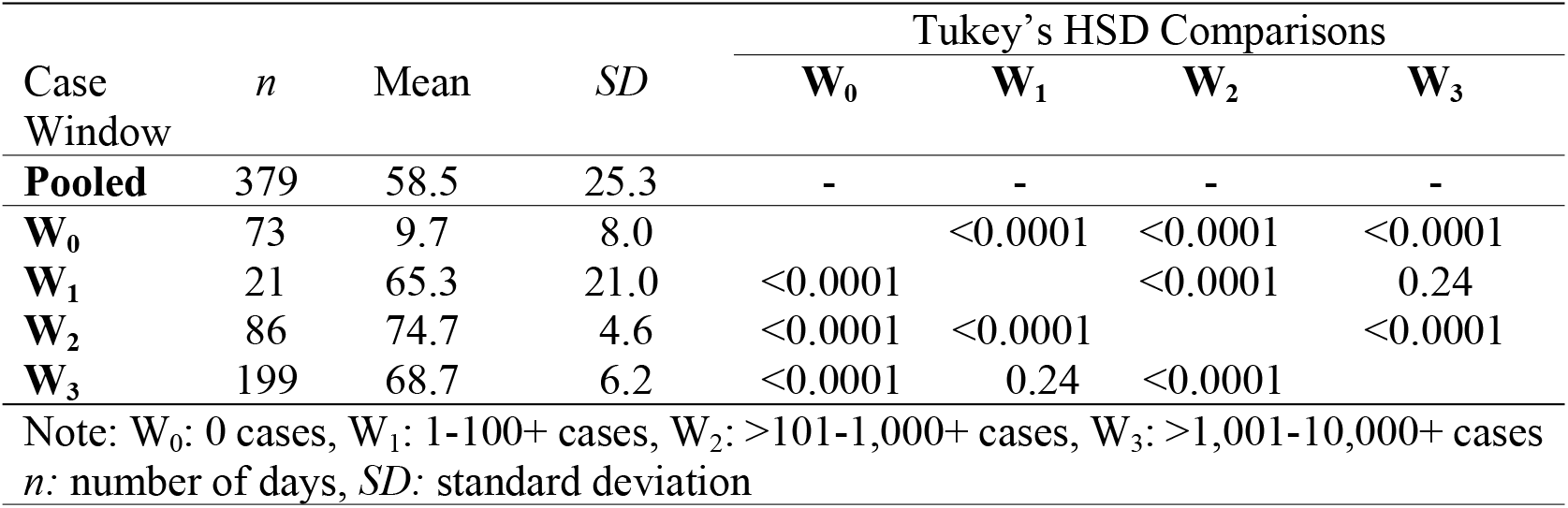
One-Way Analysis of Variance of the Containment and Health Index by Case Window, Rwanda.

**Table 3c.** One-Way Analysis of Variance of the Containment and Health Index by Case Window, Zambia.

| Case Window | *n* | Mean | *SD* | Tukey's HSD Comparisons |  |  |  |
| --- | --- | --- | --- | --- | --- | --- | --- |
| | | | | $W_0$ | $W_1$ | $W_2$ | $W_3$ |
| **Pooled** | 232 | 31.1 | 20.8 | - | - | - | - |
| **$W_0$** | 77 | 3.6 | 5.1 |  | <0.0001 | <0.0001 | <0.0001 |
| **$W_1$** | 43 | 41.7 | 13.7 | <0.0001 |  | 0.005 | 0.05 |
| **$W_2$** | 27 | 47.8 | 7.8 | <0.0001 | 0.005 |  | 0.43 |
| **$W_3$** | 85 | 45.3 | 3.3 | <0.0001 | 0.05 | 0.43 |  |

Note: $W_0$ : 0 cases, $W_1$ : 1-100+ cases, $W_2$ : >101-1,000+ cases, $W_3$ : >1,001-10,000+ cases *n*: number of days, *SD*: standard deviation

### Qualitative Findings

A total of ten (n=10) KIIs were conducted. Interviewees represented different agencies involved in the response including Ministries of Health, WHO AFRO, academic institutions, and non-governmental organizations. Several themes emerged regarding challenges in implementing NPIs from participants. The socioeconomic impact of the NPIs was a major theme. Other challenges included lack of adherence and compliance to measures and perceived severity of COVID-19 by the community. Several notable sub-themes were also identified. These are discussed below in more detail, along with illustrative statements

Nearly all participants commented on the economic hardships that certain NPIs such as lockdowns and business closures have created.

> *“… overall, the economy was affected, when businesses were closed, everywhere was closed, the economy was affected. Like we said earlier, there was also impact in accessing the services by the general population because of the movement restrictions. So, there are some of the things that were negatively impacted and has put the countries in a tight corner and made them start re-opening the economy and lifting some of those measures*.*” – Participant 1, WHO AFRO*
>
> *“Businesses really suffered. All nightclubs are completely shut down now, they’re completely out of the picture. Many restaurants shut down. And right after the lockdown was lifted, restaurants became very expensive. They had to compensate for the last one full year. That has been a challenge*.*” – Participant 3, Rwanda*

Social determinants of health such as poverty, lack of access to water, overcrowding, and food insecurity were major sub-themes that emerged. Poverty was mentioned by nearly all participants as a major challenge in both implementation and adherence to NPIs.

> *“…the challenges have been number one poverty. There are several people in Africa, Rwanda included, that depend on day-to-day wages. Take the earnings of motorcycle drivers, taxi drivers, day-laborers, farmers. Quarantine for them has been a huge, huge blow, a huge blow. Vis-a-vi other African countries did not implement quarantine. I think it was only Uganda and Kenya that did so. So that has been a big challenge*.*” – Participant 3, Rwanda*
>
> *“And then you also want to look at the fact that when you tell me wash your hands, and there’s no running water in your area, how are you going to wash your hands? Or when you say keep a safe distance and then you have a family of 6 living in a room, how are they going to keep a safe distance? And you have the whole block of single room apartments which is packed with so many people. Or in the marketplace, how do you keep distance in the market? So, some of the non-pharmaceutical interventions were not implementable because of the environment we are in*.*” – Participant 1, WHO AFRO*

An additional theme was perceived severity of disease. Several noted that media outlets, especially in the West, had emphasized that when COVID-19 reached Africa, it would create complete catastrophe. When this was not the case in the first and second wave, KII participants noted that citizens did not perceive COVID-19 to be a major threat and did not find the strict measures to be justified. KII participants mentioned that this view was further intensified by misinformation, and, in the case of Nigeria, major distrust in the government.

> *“First of all, I’m sad to say that the major challenge was the people didn’t trust the government. Two, they didn’t believe there was COVID because people were not dying because the wrong impression was created that when COVID came to Africa it was going to kill everybody, and so when they didn’t see any dead bodies then they just assumed that there was no COVID. And there was a lot of fake information and things about COVID, which people took as truth*.*” – Participant 1, Nigeria*
>
> *“…the number one challenge is the people’s perception of the disease itself because despite all the government did to make people understand how the disease is caught, you will hear people say it is a scam, that it is an attempt for government to just make money out of donors, that there’s no such disease as COVID… there was buy-in initially but because of these fear…the fear that the government cannot be trusted, because people think they had what they call “hidden agenda*.*” – Participant 2, Nigeria*

Lack of compliance and adherence to NPIs was also a major theme that emerged. In Zambia, the pandemic also coincided with a particularly tense election year, which brought its own challenges in terms of adherence to measures.

> *“… it’s been an election year for Zambia. This is unique to us so we had political gatherings that I think needed time…they didn’t really adhere to us under our recommendations, or they said in principle they would accept that, and then just went ahead and did whatever they wanted to do. So, it was rather a challenging time. And we had large gatherings, political gatherings, that was a big challenge for us to manage adherence to COVID-19 regulations, as well as ensure that there is a democratic electoral process running side by side*.*” – Participant 1, Zambia*

Enforcement was a common theme among participants. In Rwanda, enforcement was mentioned as a success several times, and it was attributed to both the malleability of the Rwandese people as a population that respects its government and strict enforcement by the government.

> *“Rwandans obey the rule of law. Obey their government. If there’s a policy, it’s respected, generally. Even without an enforcer, without the involvement of the police, of law enforcement, it is generally accepted. That’s the main thing. The second thing is they’re very strict. Walking without a mask is I think $30. I’ve seen several people getting penalized right in front of my eyes in Kigali. So, they don’t joke around. If you are caught past the curfew, you will be taken to the national stadium, you’ll be kept there throughout the night—that’s punishment in addition to $100 payment*.*” – Participant 3, Rwanda*
>
> *“Like everyone had to be home at 7pm. Right now, the time is 7:45pm and if you go on the road, you won’t find anybody. Just that move of respecting the guidelines and the level of cooperation is why I think we’re successful*.*” – Participant 1, Rwanda*
>
> *“…the enforcement piece I could say that there were radio talks about any of the measures that were put in place by the government and local entities were in charge of making sure these measures are respected by the general population*.*” – Participant 2, Rwanda*

Whereas enforcement was a success in Rwanda, enforcement was a major challenge in Nigeria and Zambia. KII participants noted that the length and protracted nature of the response have had major consequences in terms of compliance and adherence to several public health and social measures that were being implemented at various points throughout the pandemic, not only by the community but some KIIs mentioned government figures themselves were not adhering to precisely the measures they were responsible for enforcing.

> *“…there were not really [consequences for lack of adherence] …well they put it there…but complying with the law and enforcing is one thing…on a few cases here and there they did. But it didn’t last. In about a week or two everybody had forgot, and they went back…even the guy who is supposed to be enforcing it is not wearing a mask *laughs*” – Participant 1, Nigeria*
>
> *“For places of worship, they are now back to their old, crowded form despite the regulations. Because the latest law was no gathering of 50 or more people, but I was in church on Sunday, and I think we were more than 2,000 in that gathering so that’s how it is. So, there is no consistency or uniformity in the enforcement. Like every state or local government can do whatever they felt like implementing*.*”– Participant 2, Nigeria*
>
> *“…in some of the large density populations in the peri urban areas, if you went there, they would just see blatant disregard of the regulations despite posters being around or motor vehicles with the speakerphone going around announcing what to do. You would essentially see nobody wearing for instance a mask, and that just shows you the defiance levels people had, and because we didn’t really have people going around arresting you for not putting on a mask for instance, so that was some of the challenges we saw*.*” – Participant 1, Zambia*

In the opinion of KII participants, political structures and dynamics also played a factor as a barrier in the implementation of NPIs. In countries like Nigeria and Zambia, where there is a decentralized government, enforcement was much more difficult than in a country like Rwanda, which has a very centralized government.

> *“Over time, there was no consistency in the enforcement and no uniformity between States. Some States tended to take it more seriously than other States. But on the part of the Federal Government of Nigeria, the government had been the one really wielding the big stick. The States were left to do what they felt like doing because at first when the [federal] government tried to send down regulations right from the country’s capital, some State governments saw it as [an] affront to their own rights because the body governance and the president were elected so they didn’t like the idea of a [federal] government official trying to decide what happens at the State level. So, the States were left to do what they wanted to do*.*” – Participant 2, Nigeria*
>
> *“It also depends on the political structure of the country. Nigeria is a very complex country where it has federal [territories] and states. The states have very much power in terms of dealing with the local issues, so the application and implementation of these measures at the local level was a little bit not much uniform in the country. Whereby in countries like Rwanda, it’s most centralized, of course with good leadership, the implementation of these measures was very much uniform across the country*.*” – Participant 2, WHO AFRO*

## Discussion

To the best of our knowledge, this study is the first to focus on understanding the degree of implementation and facilitators and barriers to enforcement in sub-Saharan Africa. In the early stages of the pandemic, many countries in sub-Saharan Africa including Nigeria, Rwanda, and Zambia replicated NPIs, such as travel restrictions and shelter-in-place orders, as implemented by Western countries in Europe and the United States. However, approaches that were effective in countries outside of Africa were not necessarily appropriate for the African region. Given the heterogeneity in populations, health systems, and governments in the region, blanket NPI measures such as restrictions on movement proved challenging to implement. Almost three-fourths (71%) of Africans work in the informal sector and thus encountered severe economic hardship with the enforcements of lockdowns and border closures (18,27,28). With the implementation of certain measures, some countries had food shortages, social unrest, and economic instability (18,19). Economic instability was felt across the African continent; however, Zambia became the first African country to default on its Eurobond national debt during the pandemic (29). The pandemic caused the Zambian economy to enter its deepest recession in history with the economy shrinking by 4.2% in 2020 (30–32). An assessment of the Zambian economy a year into the pandemic claimed that the “recession goes beyond the containment measures (which were moderate) and reflects vulnerabilities to external shocks and unfavorable internal macroeconomic decisions, with potential long-term implications” (33).

Additionally, as key informants noted, enforcement of NPIs was met with resistance and noncompliance in countries where governmental authority was weak or contested, or misinformation was high (34–37). Similarly, physical or social distancing measures were also difficult to enforce and implement. Aside from the high population density in many communities in Africa, social interaction is a key aspect of life. In urban areas in Africa, public transportation systems are often overcrowded, dense shanty towns and informal settlements are part of the physical infrastructure, and many people do not have the luxury to self-isolate even if they are positive, as many homes face overcrowding. For example, Makoko in Lagos, Nigeria has 300,000 people whose homes are built on stilts in a lagoon (35). In rural areas, many households share sanitation facilities and have only access to water from a communal tap. Ekumah et al. (2020) used demographic and health survey data from 25 countries in sub-Saharan Africa to explore how vulnerability to COVID-19 was affected by access to basic necessities (sanitation, water, and food) within a household (38). They found that 46% of sampled households (except South Africa) lacked access to any of these three basic necessities, and only 8% had access to all three (38). Thus, physical distancing measures, including shelter-in-place measures, were unrealistic in overcrowded areas with inadequate sanitation as pointed out by KIIs.

In addition to several long-term effects of implementing NPIs, timing plays a crucial role in the implementation of NPIs. Several KIIs including those at WHO AFRO pointed out early measures undertaken by countries. However, experts may find it challenging to determine the optimal time to implement different interventions. If governments wait too long, this may lead to the proliferation of disease at a rapid rate and overwhelm health systems quickly. Consequently, the roll-out of NPIs that are too premature or uniform across an entire country may also increase the risk of a “second wave” of infections once the initial measures are halted or pandemic fatigue sets in (39,40). The implementation of NPIs, especially over a prolonged time period, can have significant detrimental consequences in terms of social and economic costs as was the case in Nigeria, Rwanda, and Zambia. While NPIs are generally effective in reducing the burden of disease and alleviating pressure on health infrastructures, studies have found that a longer duration of NPI implementation may have consequences such as increased unemployment, economic hardship, and social disruption (11). Resource-poor settings are at an especially increased risk, with some data showing income reduction as great as 70% and reduction in consumption expenditure by 30% (11,41,42). The Africa Research, Implementation Science, and Education (ARISE) Network conducted a telephone survey in Burkina Faso, Ethiopia, and Nigeria between July and November 2020 to understand COVID-19 knowledge, attitudes, practices, and their impacts on health, nutrition, and education (43,44). The education sector was profoundly affected by school closures. Food shortages and insecurity were rampant across the three sites in the study. Consumption of a range of staple foods in all three countries declined, while the prices of staples, legumes, vegetables, fruit, and animal-source foods rose (43,45). Additionally, with increased unemployment and decreased crop production, respondents described reductions in general food intake and dietary diversity (43,45).

Compliance is also a major factor in NPI implementation. NPIs are dependent on enforcement and citizens’ willingness to comply with the measure. Compliance generally wanes the longer measures are in effect (10). Poverty and economic dislocation also reduce compliance especially with NPIs focused on containment (i.e., shelter-in-place) which again was supported by KIIs in our study (46). In Rwanda, compliance with public health measures, including mask-wearing, was governed strictly by police and an anti-corona task force. Anecdotal reports detailed police pulling over cars with unmasked drivers and/or passengers, hand-washing stations and sanitizer dispensers were monitored for use before entering businesses, and arrests were made of those violating the country-wide curfew (47). The strictness with which measures were implemented in Rwanda was supported by claims made by KIIs. This was also supported by the quantitative data where the pooled average SI and CHI scores for Rwanda were much higher than that of Nigeria and Zambia. Additionally, despite a robust public information campaign to dispel misinformation, many residents in Nigeria did not initially adhere to the recommendations aimed at reducing the spread of COVID-19. This caused high tension between the civilians and armed forces who became violent when trying to enforce certain protocols (48,49). To enforce lockdown and curfew measures, policy, paramilitary, and military personnel were deployed to various areas in the country. Among the challenges in this implementation were persistent corruption and political distrust (48,49). There were also multiple reports of unlawful use of force and misconduct of the Nigerian police while enforcing COVID-19 measures (50,51). Implementation and enforcement were marred by a lack of compliance from the public, which limited the outcomes of the government response to COVID-19. An article in The Guardian described Nigerians defying the stay-at-home orders which some attributed to distrust in the government and rising reports of hunger (34). There were also reports of security operatives being susceptible to bribes from those choosing to defy lockdown measures (48). This weakened the impact of travel restrictions and lockdown measures on the slowing down of COVID-19 in the country which was also supported by KIIs.

The study has several limitations. First, given the fluidity of the COVID-19 pandemic and the time it took each of the three countries to get there, external factors such as variants, holiday season, etc., could have affected the degree of implementation. For example, Nigeria reached 10,000 cases in May 2020 while Rwanda reached that point in January 2021. The state of the pandemic and global guidance had changed significantly in between that time. There are also specific limitations in the OxCGRT dataset itself. The dataset does not measure implementation or compliance, nor does it provide subnational measures for almost all countries apart from adding a flag denoting whether the restriction was national or subnational (25). Thus, our nation-focused analysis may miss some variation of policies implemented at the sub-national level. Additionally, our sample size of KIIs is relatively small, therefore there may be other diverse opinions about what worked and what did not during NPI implementation, that were not captured here.

During pandemics, apart from effective vaccine strategies, NPIs are one of the most important tools that individuals and communities can utilize to limit disease spread and reduce deaths. In addition, the timing of NPI implementation is crucial. Delayed implementation of NPIs will lead to unchecked proliferation of disease in the community and overwhelm health systems (4,8,39). However, in addition to timing, the success of NPIs depends critically on the fidelity of implementation and the willingness of individuals to comply with the NPIs (52–55). Careful consideration of tailored NPI measures for the specific community context may lead to less resistance and improved compliance.

## Data Availability

The data that support the findings of this study are available from the corresponding author, upon reasonable request.

## References

1. Aledort JE, Lurie N, Wasserman J, Bozzette SA. Non-pharmaceutical public health interventions for pandemic influenza: an evaluation of the evidence base. BMC Public Health [Internet]. 2007 Aug 15 [cited 2021 Jul 27];7(1):208. Available from: https://doi.org/10.1186/1471-2458-7-208

2. Institute of Medicine (US) Forum on Microbial Threats. Strategies for Disease Containment [Internet]. Ethical and Legal Considerations in Mitigating Pandemic Disease: Workshop Summary. National Academies Press (US); 2007 [cited 2020 Apr 16]. Available from: https://www.ncbi.nlm.nih.gov/books/NBK54163/

3. Markel H, Lipman HB, Navarro JA, Sloan A, Michalsen JR, Stern AM, et al. Nonpharmaceutical Interventions Implemented by US Cities During the 1918-1919 Influenza Pandemic. JAMA. 2007 Aug 8;298(6):644–54.

4. Centers for Disease Control and Prevention. Nonpharmaceutical Interventions (NPIs) [Internet]. 2019 [cited 2020 Apr 16]. Available from: https://www.cdc.gov/nonpharmaceutical-interventions/index.html

5. Bo Y, Guo C, Lin C, Zeng Y, Li HB, Zhang Y, et al. Effectiveness of non-pharmaceutical interventions on COVID-19 transmission in 190 countries from 23 January to 13 April 2020. International Journal of Infectious Diseases. 2021 Jan 1;102:247–53.

6. Flaxman S, Mishra S, Gandy A, Unwin H, Coupland H, Mellan T, et al. Report 13: Estimating the number of infections and the impact of non-pharmaceutical interventions on COVID-19 in 11 European countries [Internet]. 35. 2020 Mar [cited 2021 Jul 27]. Available from: http://spiral.imperial.ac.uk/handle/10044/1/77731

7. Lai S, Ruktanonchai NW, Zhou L, Prosper O, Luo W, Floyd JR, et al. Effect of non-pharmaceutical interventions for containing the COVID-19 outbreak in China [Internet]. Infectious Diseases (except HIV/AIDS); 2020 Mar [cited 2020 Apr 16]. Available from: http://medrxiv.org/lookup/doi/10.1101/2020.03.03.20029843

8. Banholzer N, Weenen E van, Lison A, Cenedese A, Seeliger A, Kratzwald B, et al. Estimating the effects of non-pharmaceutical interventions on the number of new infections with COVID-19 during the first epidemic wave. PLOS ONE. 2021 Jun 2;16(6):e0252827.

9. Hatchett RJ, Mecher CE, Lipsitch M. Public health interventions and epidemic intensity during the 1918 influenza pandemic. Proceedings of the National Academy of Sciences. 2007;104(18):7582–7.

10. Briscese G, Lacetera N, Macis M, Tonin M. Compliance with COVID-19 social-distancing measures in Italy: the role of expectations and duration. 2020;

11. Chowdhury R, Heng K, Shawon MSR, Goh G, Okonofua D, Ochoa-Rosales C, et al. Dynamic interventions to control COVID-19 pandemic: a multivariate prediction modelling study comparing 16 worldwide countries. Eur J Epidemiol. 2020 May 1;35(5):389–99.

12. Hallas L, Hatibie A, Majumdar S, Pyarali M, Hale T. Variation in US States’ Responses to COVID-19. University of Oxford. 2021;

13. Akter S. The impact of COVID-19 related ‘stay-at-home’restrictions on food prices in Europe: findings from a preliminary analysis. Food Security. 2020;12(4):719–25.

14. Cameron-Blake E, Tatlow H, Wood A, Hale T, Kira B, Petherick A, et al. Variation in the response to COVID-19 across the four nations of the United Kingdom. Blavatnik School of Government, University of Oxford. 2020;

15. Koh WC, Naing L, Wong J. Estimating the impact of physical distancing measures in containing COVID-19: an empirical analysis. International Journal of Infectious Diseases. 2020;100:42–9.

16. Africa Centres for Disease Control and Prevention. Recommendations for Stepwise response to COVID-19 [Internet]. 2020 Mar [cited 2020 Apr 18]. Available from: https://africacdc.org/download/recommendations-for-stepwise-response-to-covid-19/

17. Ohia C, Bakarey AS, Ahmad T. COVID-19 and Nigeria: putting the realities in context. International Journal of Infectious Diseases. 2020 Jun 1;95:279–81.

18. Lancet T. COVID-19 in Africa: no room for complacency. Lancet (London, England). 2020;395(10238):1669.

19. Ihekweazu C, Agogo E. Africa’s response to COVID-19. BMC Med. 2020 Dec;18(1):151.

20. Massinga Loembé M, Tshangela A, Salyer SJ, Varma JK, Ouma AEO, Nkengasong JN. COVID-19 in Africa: the spread and response. Nat Med. 2020 Jul;26(7):999–1003.

21. Frank Hartwich, Massoud Hedeshi. COVID-19 effects in sub-Saharan Africa and what local industry and governments can do | UNIDO [Internet]. 2020 [cited 2021 Sep 10]. Available from: https://www.unido.org/news/covid-19-effects-sub-saharan-africa-and-what-local-industry-and-governments-can-do

22. Glaser BG, Strauss AL. Discovery of grounded theory: Strategies for qualitative research. Routledge; 2017.

23. QSR International. NVivo qualitative data analysis software. NVivo Release 1: QSR international Pty ltd. 2021.

24. Miller M. 2019 Novel Coronavirus COVID-19 (2019-nCoV) Data Repository: Johns Hopkins University Center for Systems Science and Engineering. Bulletin-Association of Canadian Map Libraries and Archives (ACMLA). 2020;(164):47–51.

25. Hale T, Angrist N, Goldszmidt R, Kira B, Petherick A, Phillips T, et al. A global panel database of pandemic policies (Oxford COVID-19 Government Response Tracker). Nat Hum Behav. 2021 Apr;5(4):529–38.

26. SAS Institute. SAS University Edition [Internet]. Cary, NC; 2020 [cited 2021 Jan 9]. Available from: https://support.sas.com/en/software/university-edition-support.html

27. Abdalla S, Galea S. Africa and Coronavirus—Will Lockdowns Work? | Think Global Health [Internet]. Council on Foreign Relations. 2020 [cited 2020 Apr 16]. Available from: https://www.thinkglobalhealth.org/article/africa-and-coronavirus-will-lockdowns-work

28. Nina Sun, Livio Zilli. COVID-19 Symposium: The Use of Criminal Sanctions in COVID-19 Responses – Enforcement of Public Health Measures, Part II [Internet]. Opinio Juris. 2020 [cited 2021 Sep 7]. Available from: http://opiniojuris.org/2020/04/03/covid-19-symposium-the-use-of-criminal-sanctions-in-covid-19-responses-enforcement-of-public-health-measures-part-ii/

29. Smith E. Zambia becomes Africa’s first coronavirus-era default: What happens now? [Internet]. CNBC. 2020 [cited 2021 Oct 7]. Available from: https://www.cnbc.com/2020/11/23/zambia-becomes-africas-first-coronavirus-era-default-what-happens-now.html

30. KPMG. Zambia - KPMG Global [Internet]. KPMG. 2020 [cited 2021 Sep 28]. Available from: https://home.kpmg/xx/en/home/insights/2020/04/zambia-government-and-institution-measures-in-response-to-covid.html

31. Ernst & Young. Zambia issues additional fiscal measures to mitigate the impact of COVID-19 [Internet]. 2020 [cited 2021 Sep 28]. Available from: https://taxnews.ey.com/news/2020-1113-zambia-issues-additional-fiscal-measures-to-mitigate-the-impact-of-covid-19?uAlertID=Sd%2FG8rua1oj6%2Fl58EZ2AiA%3D%3D

32. Zeidy IA. Fiscal Policy Responses Limit COVID 19’s Economic Damage. 2020;18.

33. Twivwe Siwale. One year on: Zambian economy during COVID-19 [Internet]. International Growth Centre. 2021 [cited 2021 Sep 28]. Available from: https://www.theigc.org/blog/one-year-on-zambian-economy-during-covid-19/

34. Maria Diamond, Adetayo Adeowo, Onyinye Ezeilo. Hunger obeys no order, say Nigerians defying stay-at-home directive. The Guardian Nigeria News - Nigeria and World News [Internet]. 2020 Apr 25 [cited 2021 Aug 24]; Available from: https://guardian.ng/saturday-magazine/covid-19-lockdown-hunger-obeys-no-order-say-nigerians-defying-stay-at-home-directive/

35. Noko K. In Africa, social distancing is a privilege few can afford [Internet]. [cited 2021 Sep 10]. Available from: https://www.aljazeera.com/opinions/2020/3/22/in-africa-social-distancing-is-a-privilege-few-can-afford

36. Verani A, Clodfelter C, Menon AN, Chevinsky J, Victory K, Hakim A. Social distancing policies in 22 African countries during the COVID-19 pandemic: a desk review. Pan Afr Med J. 2020 Dec 14;37(Suppl 1):46.

37. Isabel Gunther. Why social distancing is a big challenge in many African countries [Internet]. 2020 [cited 2021 Sep 10]. Available from: https://phys.org/news/2020-04-social-distancing-big-african-countries.html

38. Ekumah B, Armah FA, Yawson DO, Quansah R, Nyieku FE, Owusu SA, et al. Disparate on-site access to water, sanitation, and food storage heighten the risk of COVID-19 spread in Sub-Saharan Africa. Environ Res. 2020 Oct;189:109936.

39. Sebhatu A, Wennberg K, Arora-Jonsson S, Lindberg SI. Explaining the homogeneous diffusion of COVID-19 nonpharmaceutical interventions across heterogeneous countries. PNAS [Internet]. 2020 Sep 1 [cited 2021 Jul 27];117(35):21201–8. Available from: https://www.pnas.org/content/117/35/21201

40. Davies NG, Kucharski AJ, Eggo RM, Gimma A, Edmunds WJ, Jombart T, et al. Effects of non-pharmaceutical interventions on COVID-19 cases, deaths, and demand for hospital services in the UK: a modelling study. The Lancet Public Health. 2020;5(7):e375–85.

41. Lora Jones, Daniele Palumbo, David Brown. Coronavirus: How the pandemic has changed the world economy - BBC News [Internet]. 2021 [cited 2021 Sep 7]. Available from: https://www.bbc.com/news/business-51706225

42. Lora Jones, David Brown, Daniele Palumbo. Coronavirus: A visual guide to the economic impact - BBC News [Internet]. Diplomatic Academy. 2020 [cited 2021 Sep 7]. Available from: https://www.unic.ac.cy/da/2020/05/08/coronavirus-a-visual-guide-to-the-economic-impact-bbc-news/

43. Hamer DH. Short-term and Potentially Long-term Negative Impacts of COVID-19 in Sub-Saharan Africa: Evidence from the Africa Research, Implementation Science, and Education Network Rapid Monitoring Survey. The American Journal of Tropical Medicine and Hygiene. 2021 Aug 11;105(2):269–70.

44. Hemler EC, Korte ML, Lankoande B, Millogo O, Assefa N, Chukwu A, et al. Design and Field Methods of the ARISE Network COVID-19 Rapid Monitoring Survey. The American Journal of Tropical Medicine and Hygiene. 2021 Aug 11;105(2):310–22.

45. Madzorera I, Ismail A, Hemler EC, Korte ML, Olufemi AA, Wang D, et al. Impact of COVID-19 on Nutrition, Food Security, and Dietary Diversity and Quality in Burkina Faso, Ethiopia and Nigeria. Am J Trop Med Hyg. 2021 Jun 23;tpmd201617.

46. Wright AL, Sonin K, Driscoll J, Wilson J. Poverty and economic dislocation reduce compliance with COVID-19 shelter-in-place protocols. Journal of Economic Behavior & Organization. 2020;180:544–54.

47. Bethany Murphy. Safer in Rwanda: Other Countries Are Taking COVID-19 Seriously, and It Shows - Ms. Magazine [Internet]. 2020 [cited 2021 Sep 22]. Available from: https://msmagazine.com/2020/10/29/safer-in-rwanda-other-countries-are-taking-covid-19-seriously-and-it-shows/

48. Okolie-Osemene J. Nigeria’s Security Governance Dilemmas During the Covid-19 Crisis. Politikon. 2021 Apr 3;48(2):260–77.

49. Ezeibe CC, Ilo C, Ezeibe EN, Oguonu CN, Nwankwo NA, Ajaero CK, et al. Political distrust and the spread of COVID-19 in Nigeria. Global Public Health. 2020 Dec 1;15(12):1753–66.

50. Philip Obaji Jr. Women ‘abused’ by police enforcing COVID-19 rules in Nigeria [Internet]. 2020 [cited 2021 Oct 13]. Available from: https://www.aljazeera.com/features/2020/9/9/women-abused-by-police-enforcing-covid-19-rules-in-nigeria

51. Aborisade RA. Accounts of Unlawful Use of Force and Misconduct of the Nigerian Police in the Enforcement of COVID-19 Measures. J Police Crim Psych. 2021 Sep 1;36(3):450–62.

52. Yapi RB, Houngbedji CA, N’Guessan DKG, Dindé AO, Sanhoun AR, Amin A, et al. Knowledge, Attitudes, and Practices (KAP) Regarding the COVID-19 Outbreak in Côte d’Ivoire: Understanding the Non-Compliance of Populations with Non-Pharmaceutical Interventions. International Journal of Environmental Research and Public Health. 2021 Jan;18(9):4757.

53. Fellows IE, Slayton RB, Hakim AJ. The COVID-19 Pandemic, Community Mobility and the Effectiveness of Non-pharmaceutical Interventions: The United States of America, February to May 2020. 200712644 [q-bio, stat] [Internet]. 2020 Jul 9 [cited 2021 Jul 27]; Available from: http://arxiv.org/abs/2007.12644

54. Harling G, Gómez-Olivé FX, Tlouyamma J, Mutevedzi T, Kabudula CW, Mahlako R, et al. Protective behaviours and secondary harms from non-pharmaceutical interventions during the COVID-19 epidemic in South Africa: a multisite prospective longitudinal study. medRxiv [Internet]. [cited 2021 Jul 27]; Available from: https://www.ncbi.nlm.nih.gov/pmc/articles/PMC7668759/

55. Kantor BN, Kantor J. Non-pharmaceutical Interventions for Pandemic COVID-19: A Cross-Sectional Investigation of US General Public Beliefs, Attitudes, and Actions. Frontiers in Medicine. 2020;7:384.

